# Co-Creation of Activity Spaces: A study protocol for the evaluation of co-created local community interventions to support activity participation in youth across municipalities in Eastern Norway

**DOI:** 10.1101/2025.10.21.25338441

**Authors:** Emma Charlott Andersson Nordbø, Camilla Ihlebæk, Ellinor Moe, Guro Amundsen, Elling Bere, Charlotte Pawlowski, Jasper Schipperijn, Ingeborg Pedersen

## Abstract

**Background:** Important gaps remain in our understanding of the complexity and impact of creating activity spaces for youth as a public health measure. This protocol outlines the study design, cases, and methods for the Co-Creation of Activity Spaces study, which evaluates the development, implementation, and impact of activity spaces in local communities in semi-urban and rural municipalities in Eastern Norway.

**Methods:** The activity spaces are developed within the context of the development project *EMO (A Fun Detour)* through co-creation processes with multiple stakeholders, including local youths. The evaluation study takes a pragmatic mixed-method approach, using a quasi-experimental design. It consists of three sub-studies (i.e., Process, Places and People) targeting the co-creation processes, the use of the activity spaces, and the potential impact for local youths. To gain knowledge of the processes, focus group interviews and concept mapping with involved stakeholders will be conducted. Usage of the activity spaces will be assessed through a monitoring system based on AI-technology and on-site observations. Finally, the impact will be examined through repeated cross-sectional surveys conducted before and after the establishment of the activity spaces, along with qualitative data from walk-along interviews and photovoice studies with local youths.

**Discussion:** Through its focus on semi-urban and rural contexts and by using a novel method to capture long-term use of the activity spaces, the study will contribute new knowledge. The results will offer valuable insights into the co-creation processes, the evolving use of the newly constructed activity spaces, and their impact on local youths’ engagement in activities. This information can guide future projects aiming to establish youth activity spaces in both semi-urban and rural local communities.

## Background

Repeatedly, studies have underscored the importance of maintaining regular physical activity [1-3] and being involved in social networks and activities [4-6] for health and wellbeing in youth. Yet, a substantial number of youths worldwide do not engage in sufficient physical activity, and the activity level is declining [7]. This trend is observed across all Western countries [7], including Norway, where the proportion of inactive children has increased along with significant declines in activity levels throughout adolescence [8]. Moreover, estimates show that up to 15% of youth worldwide experience social isolation and loneliness [9], and in Norway, there has been a steady increase in loneliness among Norwegian adolescents since 2014 [10], which exacerbated during the COVID-19 pandemic [11].

From this, it is evident that promoting activity participation remains critical for the health and wellbeing of youth, and efforts to encourage active living are therefore a public health priority [12]. With the recognition of neighborhoods as key contexts for health-promoting actions aimed at supporting active living [13], attention to the design of the neighborhood built environment has increased over the last decade [14, 15]. A well-designed built environment has the potential to facilitate participation in both physical and social activities [14, 16, 17], and different public health measures that involve modifications to the built environment, like public open space interventions, have been implemented [18-22].

Implementing such local community interventions has the potential to address major challenges, such as inactivity, loneliness, and social inequalities in health [18, 23, 24]. Importantly, the development and implementation of new outdoor activity spaces as a public health measure are context dependent and complex [25]. Nearly 75% of all settlements in Norway are small semi-urban settlements, of which many are situated in rural municipalities. Additionally, about 17% of the population resides in even smaller rural settlements [26]. This stresses the significance of developing targeted and attractive activity spaces in rural municipalities. However, when designing neighborhood interventions in semi-urban and rural areas, findings from urban studies might not be sufficient or useful. Therefore, we need more knowledge on how to develop activity spaces for youths residing outside the urban context. Additionally, research examining the potential benefits of such interventions in rural areas is warranted [27].

More research investigating the complexity of creating activity spaces, from idea to implementation, including long-term impact, is also required to comprehensively evaluate the impact of such a public health measure [18, 22, 28]. Particularly, research paying attention to the experiences of different stakeholders involved in the planning process is requested [29, 30]. One increasingly recommended approach for processes at the local level is co-creation [31]. Co-creation describes a way of understanding complex problems and designing and evaluating contextually relevant solutions by engaging diverse stakeholders in the process [32]. Few studies have examined different stakeholders’ experiences of participating in co-creation processes taking place in the public sector. Moreover, co-creation often poses challenges to the stakeholders involved [31, 33], and we need more research considering the opportunities and challenges involved when co-creating public health interventions within municipalities. Another important gap of knowledge is to understand how and by whom the activity spaces are used. This includes assessments of individual and contextual level factors to understand barriers and facilitators for use [28, 34], and usage over time must be captured by long-term follow-up [28, 30]. Lastly, studies should assess if the activity spaces influence youths’ participation in physical and social activity.

In response to the need for addressing inactivity and loneliness among youth in Norway, the *EMO – A Fun Detour* project was established in 2020. The project aimed to develop innovative activity spaces in both semi-urban and rural local communities across Eastern Norway, and six municipalities were involved in co-creation processes to develop tailored activity spaces for self-organized physical activity (see project website: https://tverga.no/emo/). The *EMO – A fun Detour* project was inspired by a similar development project conducted in Denmark [35]. Comprehensive evaluations of such projects are needed [20, 22, 36], and as highlighted above, several gaps remain to be addressed to fully understand the complexity and impact of creating activity spaces for youth as a public health measure. Therefore, we designed the *Co-Creation of Activity Spaces* study to evaluate the development and implementation of activity spaces in local communities across semi-urban and rural municipalities in Eastern Norway. In line with the identified knowledge gaps, the study focuses on examining the co-creation processes, use of the established activity spaces, and the potential benefits for local youths living nearby the activity spaces. The aim of this protocol is to present the study design, cases, methods, and measurements of the evaluation study.

## Methods

### Context

The evaluation study described in this protocol is set in the context of the Norwegian development project called *EMO – A Fun Detour*, which aimed to establish innovative activity spaces in local communities. The non-governmental organization (NGO) Tverga (www.tverga.no), which serves as the national resource center for self-organized sports and physical activity in Norway, was the initiator and project owner. The NGO received approximately € 1.306.500 from The Savings Bank Foundation DNB to cover process and planning costs and to partially fund the local activity spaces. The development project was organized as an open competition for municipalities in Eastern Norway. The NGO advisors were responsible for organizing the competition and development process, in collaboration with the consultants from Asplan Viak, which is one of the leading companies in planning, architecture and engineering in Norway, and researchers from the Norwegian University of Life Sciences (NMBU).

### Competition and case selection process

In October 2020, all 121 municipalities in Eastern Norway received an invitation to submit a proposal for developing an innovative activity space. The invitation specified that municipalities had to establish an interdisciplinary project group, including municipal staff from relevant departments such as public health, education, parks, and recreation, to qualify for participation. Additionally, the proposal needed to include a needs assessment justifying the local need for an activity space and identifying the population target group that would benefit from an activity space. Out of the 38 submitted proposals, 10 cases were selected by the partners in the *EMO – A Fun Detour* project (Tverga, Asplan Viak, and NMBU) and a representative from The Savings Bank Foundation DNB based on a rating (from 1-5) on five focus areas from the application (i.e., justification, location, potential, innovation, and uniqueness), as well as the motivation and ambition of the municipal project group. The 10 selected municipalities were given the opportunity to further develop their plans, and during spring 2021, the NGO advisors organized two workshops for the municipalities to support the development of their specific plans. During the workshops, the municipalities received extensive feedback on their draft plans from the consultants. In April 2021, the 10 municipalities submitted and presented their revised plans. Based on the revised plans, an evaluation panel comprising of six experts from the NGO and the consultants finally selected seven municipalities. The selection was based on each municipality’s development of their plan in relation to the five focus areas (i.e., justification, location, potential, innovation, and uniqueness) in addition to the following criteria: inclusiveness (i.e., degree and content of public participation), political anchoring, and progress plans. Each of the municipalities selected at this final stage received approximately € 4.600 to support the completion of their plans. Between May and December 2021, the 7 selected municipalities refined their plans under supervision of the NGO advisors and the consultants. During this period, one of the municipalities had to withdraw from the project, due to a lack of progress with their plan for the activity space. The six remaining municipalities presented their final plans in January 2022 and received funding from the NGO (between €90.000 – 175.000) to realize their plans.

### Cases

All six municipalities aimed to create innovative activity spaces that offer low-threshold opportunities for activities in local communities, with youth (10-15-year-olds) as the main target group. The activity spaces are located in municipalities of varying centrality and population size in rural and semi-urban local communities in Eastern Norway (see Figure 1). The development of the first activity space started in autumn 2022, and all activity spaces are projected to be completed by autumn 2025. Table 1 provides a brief overview of each case, including the intended target group, expected outcomes, and intervention elements. Further details about the municipalities and the activity spaces are provided in the supplementary material (see S1 – S6).

**Table 1.** An overview of the six cases included in the evaluation study.

| **Case** | **Target group** | **Expected outcomes/aim** | **Intervention elements** |
| --- | --- | --- | --- |
| Gjøvik | Youth with a special attention to girls | Provide a space for having fun and being social, with predominantly low-intensity activities that do not necessitate traditional sport skills. | A space divided into four zones, each serving a specific purpose: a relaxation area, a cool hangout spot, a low heart rate zone and an area for fine motor activities. The elements and activities within these zones include frisbee golf, table tennis, spike ball, a designated campfire area and a net for chilling. |
| Larvik | Youth | Provide a year-round activity space with a vibrant environment where youth can engage physically, socially, and digitally through game-based activities. | Twelve game and activity zones will be created, including a mobile app where youth can create their own avatars. Young gamers can earn “parcoins” by participating in physical challenges across different zones, such as “the arena”, featuring various ball fields, workout furniture, and interactive play equipment, as well an amphitheatre stage and a DJ booth. |
| Nordre-Follo | Youth | Develop a year-round activity space along parts of a canal, serving both as a local attraction and a regional destination. The activity space will seamlessly blend with nature and connect the canal to the nearby lake. | A canal path adorned with small information signs about the nature, plant, and animal life in the area. At each end of the canal there will be a harbour. One of the harbours will house a café featuring a simple roof, outdoor and there be a socialization zone with a semi-open hut with benches and hammocks. |
| Trysil | Youth | Provide an exciting space for both physical and social activity by creating a stronger connection between the main center and the river. | A solid steel pipeline that serves as an obstacle course near the water, accompanied by swimming area, and hammocks and nets for socializing. |
| Vestby | Youth with a special attention to girls | Establish an attractive, social, and inclusive activity space for girls who want to roller skate. | A circular roller-skating arena with concrete surface. The space will also consist of asphalt paths, two circle shaped stations with mini ramps and benches, a self-service equipment shed, tables and benches outside the shed, and other hangout areas. The roller-skating arena and its surroundings will be decorated with diverse artistic elements. |
| Vinje | Youth aged 9-16 years | Provide a year-round activity space for youth to meet and engage in physical and social activities. | A diverse activity space including a pump track, a bike park, skill trails, a combined jogging and walking path through the forest, and an elevated floating path among the treetops. |

**Fig. 1.**
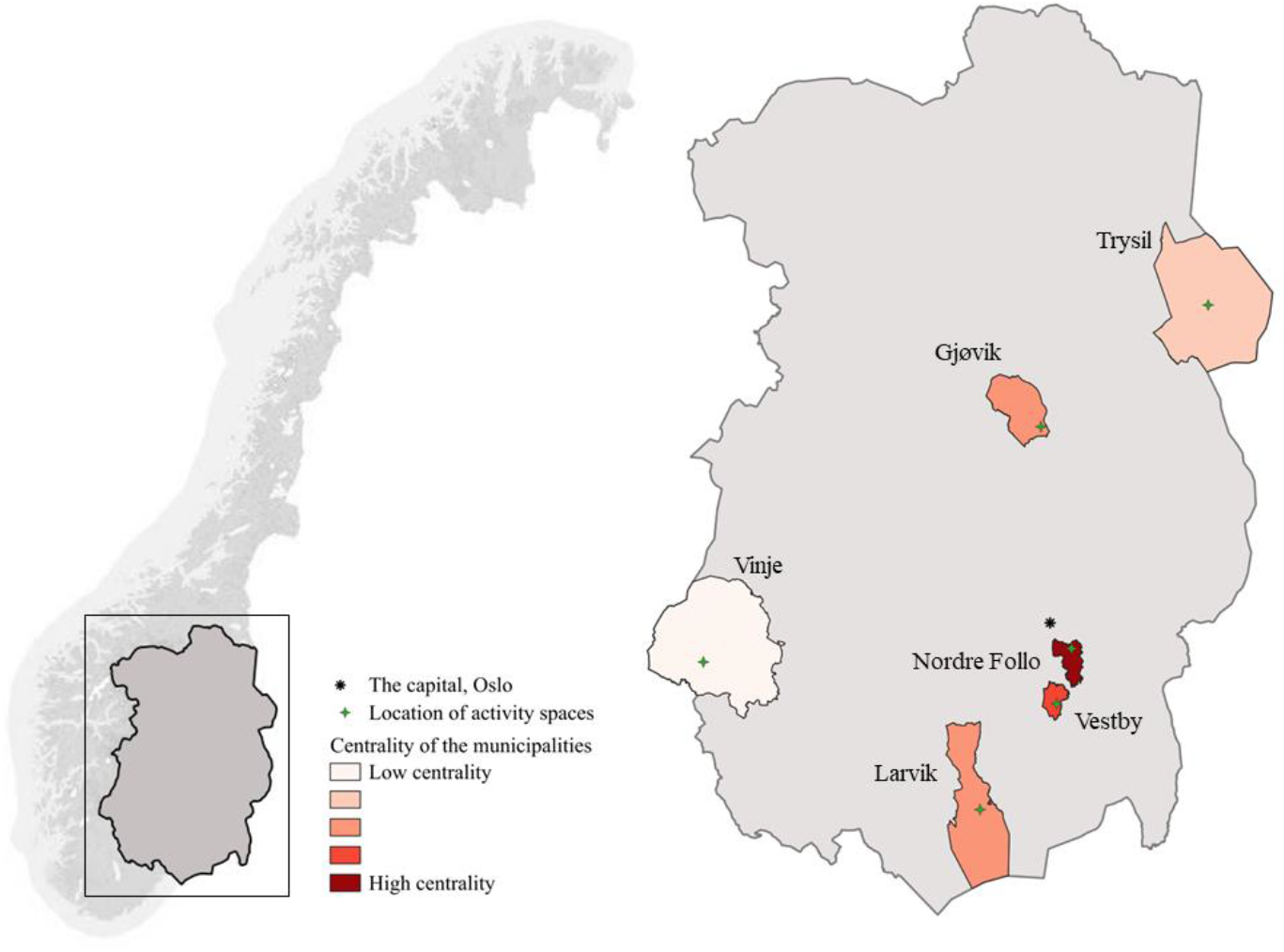
Map of Norway displaying the location of each municipality, categorized according to centrality, and their respective activity spaces within Eastern Norway.

### Research design

The current evaluation study (*Co-Creation of Activity Spaces*) employs a pragmatic mixed-method approach and a quasi-experimental design to address the complex nature of the development and implementation of the activity spaces. The study comprises three sub-studies, each with a distinct focus. The sub-study *«Process»* examines the planning and development of the activity spaces in the six local communities. The sub-study *«Places»* investigates the neighborhood contextual factors, as well as the content and use of the activity spaces. The third sub-study *«People»* centers on the inhabitants and the users of the activity spaces. Figure 2 illustrates the timeline for the development project (*EMO – A Fun Detour*) and the evaluation study (*Co-Creation of Activity Spaces*) with its three sub-studies and data collections.

**Figure 2.**
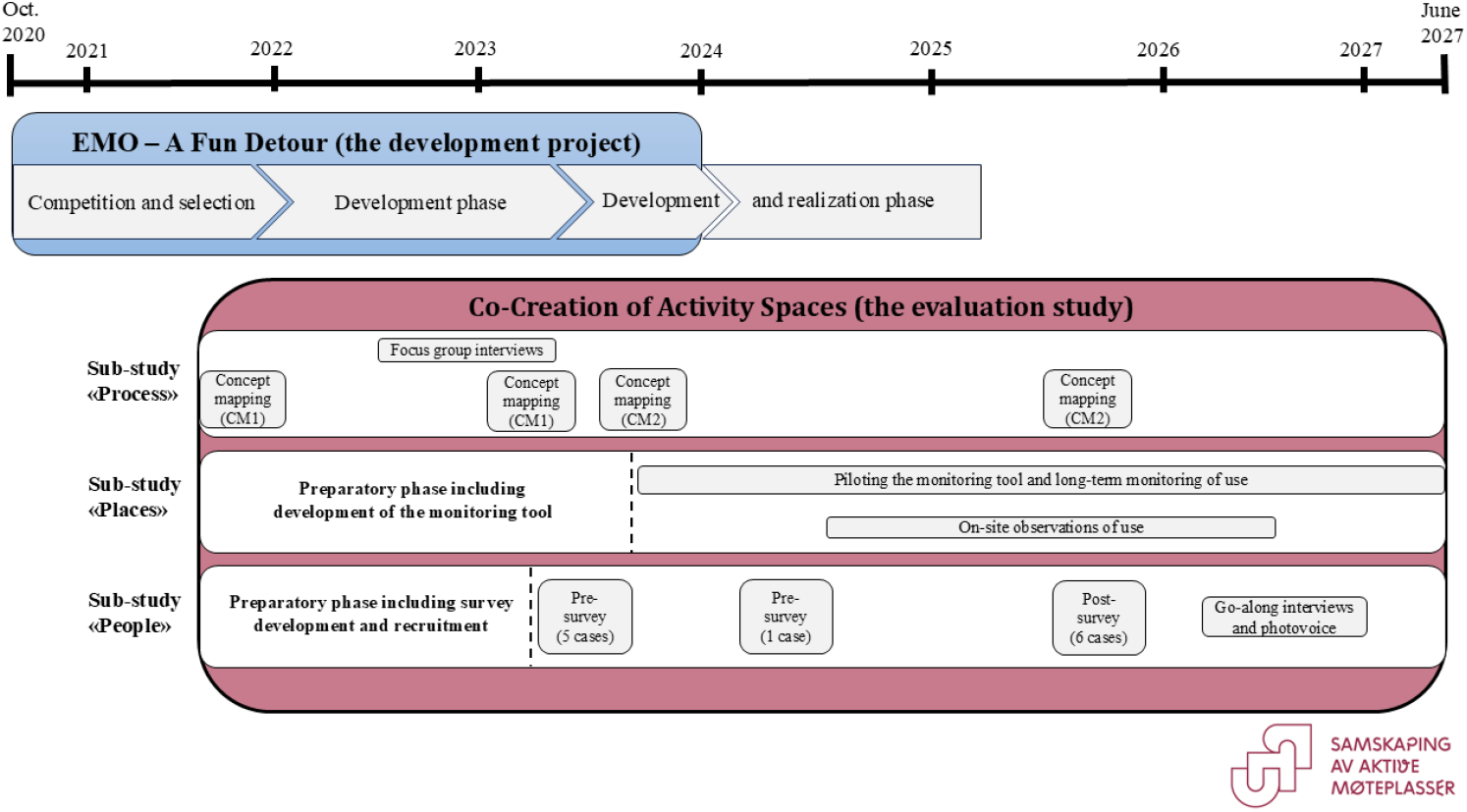
Illustration of the development project (*EMO – A Fun Detour)* and the evaluation study (*Co-Creation of Activity Spaces)*, including timeline and data collections.

### Research team

The research team includes a group of national and international researchers with different disciplinary backgrounds from the Norwegian University of Life Sciences (NMBU), Norwegian Institute of Public Health (NIPH), and University of Southern Denmark (SDU).

The team brings together and combines expertise and research competence within planning, public participation, landscape architecture, physical activity, public health science, spatial analysis, intervention studies, and active living. To maintain close links with the development project, the research team will regularly meet the NGO advisors and the consultants, who are also involved as partners in the *Co-Creation of Activity Spaces* study.

### Sub-study «Process»

The primary focus of this sub-study is to investigate how involved stakeholders experience the process of establishing activity spaces within the frames of the development project. The study aims to gain in-depth knowledge of 1) stakeholders’ experiences of co-creating activity spaces, 2) how roles and power dynamics in the co-creation process are perceived by the stakeholders, and 3) what the stakeholders consider as important factors for using and for realizing the spaces.

To obtain empirical data on stakeholders’ experiences of co-creating activity spaces, and how existing roles and power dynamics in the process are perceived, group interviews will be conducted (Figure 2). All stakeholders, including the six municipal project groups, youth involved in the co-creation within each municipality, the NGO advisors, and consultants, will be invited to participate in the interviews. For participant recruitment, municipal stakeholders will be reached through project managers in each municipality, who will also provide contact information for the youth involved in the planning of each activity space. In cases where the schools will be used as co-creation arenas, we will recruit youth through the schools. The NGO advisors and consultants will be reached through our contact person in each organization. The interviews with the municipal stakeholders and adolescents will be conducted at a suitable location in the respective municipality. Interviews with the NGO advisors and consultants will occur at their offices or other suitable meeting facilities. The interview data will be analyzed using systematic text condensation [37]. The interviews were conducted between October 2022 and March 2023.

The participatory mixed-method concept mapping (CM) will be used to obtain knowledge on what stakeholders consider as important factors for use (CM1, Figure 2) and realization (CM2, Figure 2) of the activity spaces [38]. In the first CM, all stakeholder groups involved in the development project were invited to participate, while only stakeholders in the municipal project groups were invited to take part in the second CM. The data will be collected in two steps. First, two group brainstorming sessions will be carried out at different time points during the development process. The goal of the first brainstorming session is to gather ideas, experiences, and attitudes related to the use of the activity spaces (CM1). Written materials of ideas gathered through public participation with youth will supplement this brainstorming session. The second brainstorming session will address factors related to realization of the activity spaces (CM2). In step two, participants will individually sort the collected statements from step one into themes based on their assessment of how the meaning of the statements relate to each other. Additionally, the participants will rate the importance and feasibility of each statement for use and realization of the activity spaces. All stakeholders involved in the development project will be invited to participate in step two of CM1. To recruit participants for step two of CM2, municipal employees will be reached through the project managers in each municipality. A quantitative analysis will be carried out using the software Groupwisdom™ © [39] on data from step two for both CM1 and CM2 to obtain an overview of factors and mechanisms related to using and realizing the activity spaces. Data collection for CM1 took place in November 2021 and January 2023. The first round of data collection for CM2 was conducted in October 2023, with the second round scheduled for September and October 2025.

### Sub-study «Places»

This sub-study aims to achieve two objectives. First, it seeks to assess long-term usage of the developed activity spaces. Second, it aims to identify distinct user groups and examine their activity patterns within these spaces. To facilitate a comprehensive long-term assessment, the two activity spaces that are the first to open to public use will be chosen. This will ensure that the assessment spans the maximum possible timeline. To provide knowledge on the user groups and their activity patterns, the same two cases will be selected for validity and complementarity purposes. This will allow us to elucidate and elaborate upon the findings derived from the various research methods that will be applied for collecting data [40].

Longitudinal data on usage will be collected utilizing camera monitoring based on AI-technology. As soon as the activity spaces are opened, a camera will be installed at each of the selected cases. The camera has a tiny built-in computer with an embedded AI-solution that allows us to quantify moving objects directly from a video image flow. The AI-solution reads approximately 30 images per second, and moving objects are continuously detected in each image. After detection is completed, video images are deleted every second and data is automatically transferred to Microsoft Power BI that manages big data using AI-capabilities. The data will be linked to daily weather data from the Norwegian Centre for Climate Services and the Norwegian Meteorological Institute. Depending on when the projects are completed, the monitoring will last for up to three years after the activity spaces are opened to the public. Cameras were installed in the reindeer park in Vinje municipality in October 2023 and at the Trysil knot in Trysil municipality in September 2024. These activity spaces will be continuously assessed from the time of camera installation until the evaluation study concludes in June 2027 (Figure 2). We will begin processing and analyzing the monitoring data in autumn 2025. Through the camera monitoring, we will obtain measurements on daily and hourly users at the activity space and within specific zones of the space, as well as movement tracks (e.g., directions of movement). Daily activity monitoring data will be downloaded and recorded from the Microsoft Power BI platform. The data will be processed and analyzed using statistical software to examine differences in usage across weekdays and weekends, months, seasons, and according to weather conditions.

On-site observation will be used to gather detailed data on user groups, the users’ activity behaviors and activity level, and the environmental context where the activities occur. We will take advantage of existing validated systematic observational tools (e.g., SOPARC, SOPARNA) [41, 42]. To provide a robust assessment of user groups and their activity behavior, we will follow an observation schedule where observations are conducted 4 times/day over three to four days, including both weekdays and weekends [42, 43]. For each case, three on-site observations will take place throughout the year (i.e., early autumn, winter, and late spring) to capture seasonal variations (Figure 2). All users will be counted by gender (female or male), age group (child, teen, adult, or senior), type of activity (e.g., fitness activities, ball sports, active play, and recreation) and activity level (sedentary, walking, or vigorous). This data will be collected for each activity space as a whole and for each specific installation in these spaces. We will also record weather data, and the characteristics of each activity space will be described (e.g., accessibility, equipment, and lighting). On-site observations were conducted at the Trysil knot in October 2024 and March 2025. Additional data collection will take place in autumn 2025 and spring 2026. The observational data will be analyzed using descriptive statistics and chi-squared tests.

### Sub-study «People»

This sub-study has two key aims. First, it aims to examine associations between the neighborhood environment and youth participation in physical and social activities. The second aim is to assess the impact of creating tailored activity spaces in local communities on youths’ engagement in both physical and social activities, while also considering sociodemographic and contextual factors, as well as potential facilitators and barriers for use of the activity spaces.

Quantitative data will be collected through repeated cross-sectional surveys among youth that will be carried out before and after the establishment of the activity spaces (Figure 2). To reach the target group, youth aged 10–16 years residing in the six case municipalities will be recruited through the primary and lower secondary schools. We will employ two sampling strategies. To ensure the inclusion of youth living both close by and far from the activity spaces, representative sampling will be conducted by inviting youth from 5^th^ to 10^th^ grade at all schools in the municipalities. In cases where this approach is not feasible, purposeful sampling will be used to recruit youth from schools located reasonably close to the activity spaces, as they are more likely to use the spaces as part of their everyday environment.

We will use a self-developed questionnaire based on questions from Ungdata (https://www.ungdata.no/english/), which is national data collection scheme that surveys youth at the municipal level in Norway. This allows us to compare our results with existing national and regional data. The questionnaire will cover sociodemographic information, participation in various activities, including physical, social, organized, and self-organized activities during leisure time, perceptions about the neighbourhood, and transport habits. We will also collect the residential address of each participant to link the survey data to neighborhood contextual variables surrounding the participants’ home. These neighborhood variables will be computed using spatial data from the Norwegian Mapping Authorities and Open Street Map within a geographical information system (GIS) software. To compare results before and after establishment, both the pre- and post-survey will contain questions about the specific area where the activity spaces will be developed, including use and satisfaction. Additionally, the post-survey will inquire about motivations and barriers for use.

Based on the project timeline for each municipality, pre-survey data were collected between March and June 2023 (for five municipalities) and April 2024 (for one municipality). Post-survey data will be collected in September and October 2025 (Figure 2). Both the pre- and post-survey will be conducted during a school lesson using youths’ own digital devices through *Nettskjema*, a web-based survey tool that allows for creating and managing surveys, as well as collecting data in compliance with data protection regulations. The pre-survey data will be used to address the first aim, while the post-survey data will address usage of activity spaces according to sociodemographic and contextual factors, as well as barriers and facilitators for use. Additionally, data from both the pre- and post-survey will be used to investigate changes in youths’ activity pattern after development of the activity spaces. The survey data will be processed and analyzed using statistical software.

To gain in-depth knowledge of what youth perceive as facilitators or barriers for use, as well as unintentional consequences of developing the activity spaces, go-along interviews [44] and photovoice studies [45] will be conducted (Figure 2). The go-along interviews will involve accompanying the individual informants during their activities at the developed activity spaces. Participants will be recruited from schools located within a decent distance from the activity spaces in two or three of the participating municipalities. The data will be analyzed using systematic text condensation [37] and “The interpretive engagement” framework will be used to interpret the pictures [46].

## Discussion

The Co-Creation of Activity Spaces study will address several research gaps by evaluating the development and impact of co-created activity spaces for youth in both semi-urban and rural areas in six Norwegian municipalities. Given the complex and context-dependent nature of such public health interventions [25], we made several deliberate choices when designing the evaluation study to enhance its robustness.

A complex evaluation design requires the involvement of many different participants. One particular strength of this study is that it is planned and developed in collaboration between NGO advisors, consultants within architecture and construction and researchers from multiple fields, each using their core competences to share ideas and set up the evaluation. Research has shown that the more design and planning experiences are shared and have crossed professional barriers, participants are more likely to learn, generate valuable ideas, increase quality and flexibility, improve efficiency, and optimize resource utilization [47].

Another key strength of the project is the long timeframe, spanning from autumn 2021 to June 2027. The activity spaces will be planned, developed, and evaluated in real life settings and systems involving multiple stakeholders. These stakeholders may respond in unpredictable ways, and the progress of the planning and construction in the municipalities can be delayed. However, the long timeframe of the evaluation study offers flexibility and allows us to be responsive to unforeseen circumstances and delays. Ultimately, this provides a solid basis for conducting a comprehensive assessment of the impact of developing activity spaces for youth. Still, we must remain aware that delays can affect the timespan between intervention completion and follow-up, which may vary across municipalities.

Further, combining a process evaluation with an effect evaluation, and collecting different types of data through complementary methods, represent strengths of this study evaluating a complex intervention [48]. We also opted for a cross-sectional design to avoid the age-dependent decline in physical activity [49]. To fully comprehend activity space usage, long-term follow-up is essential [28, 30]. We will collect longitudinal data by developing and applying an innovative monitoring system based on AI-technology. The longitudinal data on usage will be linked to daily weather data, allowing us to explore usage patterns across weekdays and weekends, months, seasons, and according to weather conditions. This represents a strength and a unique contribution of this evaluation study.

It is also important to acknowledge some of the limitations of the evaluation design. The study employs a quasi-experimental cross-sectional design, studying the same age group at two distinct time points. This means that we will compare two different samples of youth. In contrast to the randomized controlled trial (RCT) design, where participants are randomly selected, the municipalities receiving the intervention will be purposefully selected. In principle, not using RCT design reduces the internal validity of a study [50]. This means that potential changes in youth health behavior may not be entirely attributed to the intervention. However, a traditional RCT design is not appropriate for studying a project co-created within complex systems as the design lack the ability to measure and understand the process and contextual effects, including unintended effects on other parts of the system [51].

We intentionally excluded municipalities from the development and planning of the evaluation study. This decision was taken because the final six municipalities chosen for intervention were not determined at the time of developing the evaluation protocol. Some have argued that attempts to turn practitioners, such as municipal employees, into coresearchers with responsibility to balance rigor and practical relevance are overvalued in the academic discourse [52]. Nevertheless, we recognize the importance of collaborating with the selected municipalities during data collection (e.g., involving school managers) to ensure a high number of youths from the local schools in our questionnaire measurements.

## Conclusion

This study protocol has outlined how we will evaluate the process and impact of the *EMO – A Fun Detour* project using a multi-method design that includes three linked but independent sub-studies (i.e., Process, Places, and People). The results will provide unique insights into the co-creation processes, how the youth experience and use the newly built activity spaces over time and if the spaces influence the youths’ engagement in physical and social activities. This knowledge can guide future projects aiming to establish youth activity spaces in both semi-urban and rural local communities.

## Data Availability

All data produced in the present study are available upon reasonable request to the authors, but restrictions apply to the availability due to agreements and approvals involving data security for participants from the Norwegian Agency for Shared Services in Education and Research.

## Abbreviations

AI: Artificial intelligence
CM: Concept mapping
GIS: Geographical information systems
NGO: Non-governmental organization
NIPH: Norwegian Institute of Public Health
NMBU: Norwegian University of Life Sciences
SDU: University of Southern Denmark
SOPARC: System for Observing Play and Recreation in Communities
SOPARNA: System for Observing Physical Activity and Recreation in Natural Areas
RCT: Randomized controlled trial

## Declarations

### Ethics approval and consent to participate

The Co-Creation of Activity Spaces study will be carried out according to laws, regulations, and the principles in the Helsinki declaration where respect for human dignity and personal integrity are emphasized. All data from participants will be based on written informed consent (written parental consent for youth aged below 16 years) and information to participants, including the purpose of the project, details about methods, risks, and the participants’ right to withdraw from their participation in the study. Privacy and integrity of personal information will be ensured by following GDPR regulations, and personal confidentiality will be guaranteed all participants. The three sub-studies have been notified to the Norwegian Agency for Shared Services in Education and Research (ref. no.: 616232, 535488, and 784252).

## Acknowledgements

We express our gratitude to all the participating municipalities, especially the municipal project groups, for their active involvement in the co-creation process and their collaborative support throughout all phases of data collection. Special thanks are also due to our project partners and collaborators at Tverga and Asplan Viak. Lastly, we thank the school principals, teachers, the participating youth and their families for their valuable contributions and engagement in the evaluation study.

