## Supplementary material for "Co-Creation of Activity Spaces: A study protocol for the evaluation of co-created local community interventions to support activity participation in youth across municipalities in Eastern Norway"

This supplementary information contains the following:

- Supplementary information S1. The reindeer park in Vinje
    - **Figure S1.** Overview of Vinje municipality and the location of the activity space.
- Supplementary information S2. The Trysil knot
    - **Figure S2.** Overview of Trysil municipality and the location of the activity space.
- Supplementary information S3. The pool park in Gjøvik
    - **Figure S3.** Overview of Gjøvik municipality and the location of the activity space.
- Supplementary information S4. The Parcade in Larvik
    - **Figure S4.** Overview of Larvik municipality and the location of the activity space.
- Supplementary information S5. The Bruer park in Vestby
    - **Figure S5.** Overview of Vestby municipality and the location of the activity space.
- Supplementary information S6. Little Venice of Norway, the Siggerud canal
    - **Figure S6.** Overview of Nordre Follo municipality and the location of the activity space.

### Supplementary information S1. The reindeer park in Vinje

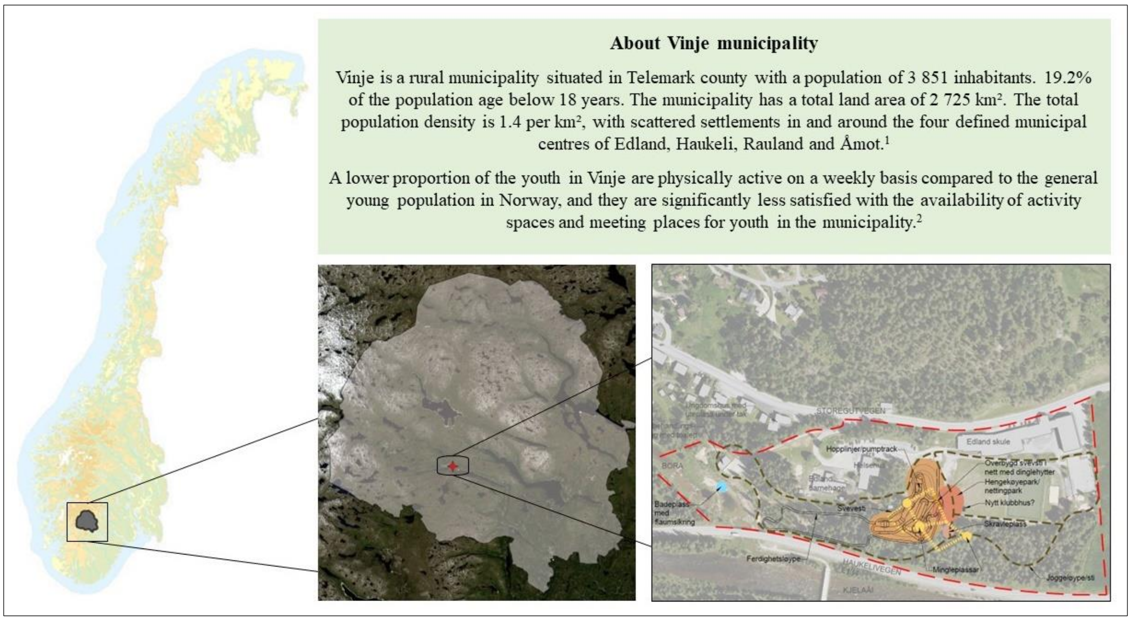


[1] Statistics Norway (2024). *11342: Population and area, by region, contents, and year.* [Online]. Available from: https://www.ssb.no/en/statbank/table/11342/tableViewLayout1/ (downloaded 31st May 2024).
[2] Norwegian Institute of Public Health (2024). *Youth profiles for municipalities and districts.* [Online]. Available from: https://www.fhi.no/op/oppvekstprofiler/ (downloaded 31st May 2024)

**Figure S1.** Overview of Vinje municipality and the location of the activity space.

**Target group, location, and aim**

Vinje municipality aims to create a year-round activity space in the local center of Edland. This activity space will be adjacent to facilities such as a kindergarten, school, health center, youth center, and soccer, handball, and volleyball fields. See Figure S1 for a general overview of the municipality.

The activity space intends to provide a meeting place and activities that engage and inspire youth aged 9-16 years to participate in physical and social activities.

**Concept and design elements**

To achieve the aim, a pump track, a bike park, skill trails, a combined jogging and walking path through the forest, and an elevated floating path among the treetops will be installed. The flood embankment surrounding an artificial pond will be designed to encourage activity and play, while the pond itself will serve as a swimming area with a designated small dock. Additional elements include several semi-open huts with campfires that are suitable for overnight stays and other spots to socialize and chill, like floating huts in the treetops, hammocks, and nets. The design of the play equipment will incorporate the distinctive antlers of the reindeer. The elements will be developed in four construction stages and designed to be universally accessible. Furthermore, the activity space will adapt to the changing seasons. For instance, the area will cater to regular cycling during the summer and fat biking during the winter, while the pond, initially set up for swimming in the summer, will transform into an ice-skating area in the winter.

**Stakeholders involved in the project**

Interdisciplinary municipal project group, local youth, external consultants, and the inhabitants.

### Supplementary information S2. The Trysil knot

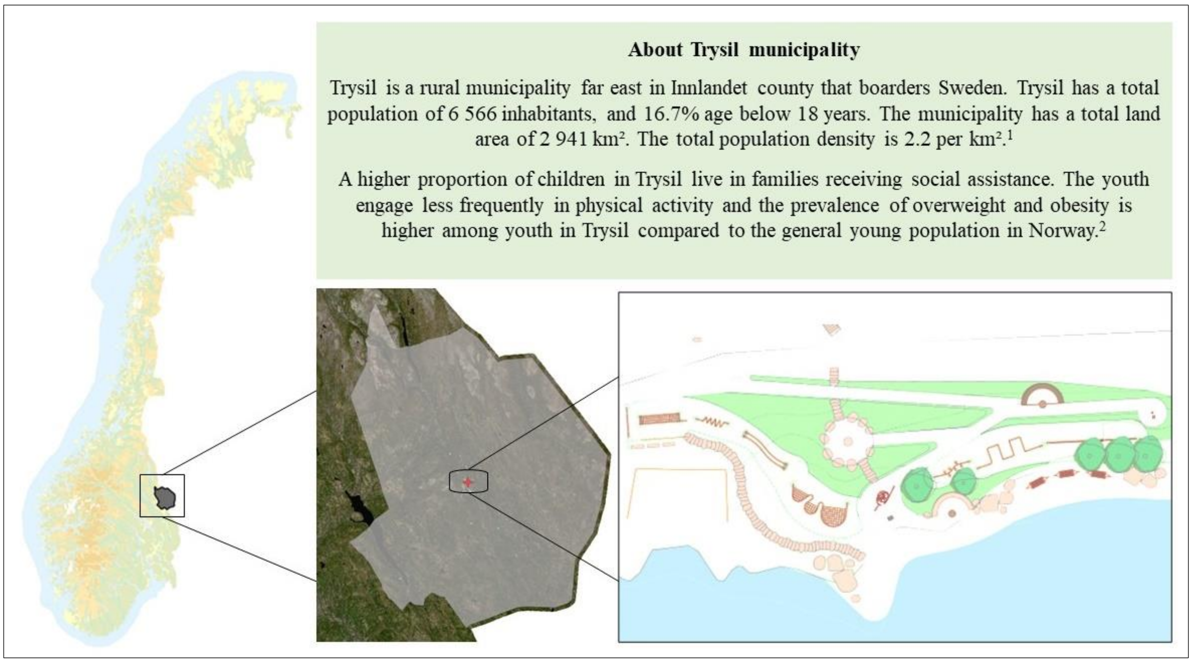


[1] Statistics Norway (2024). *11342: Population and area, by region, contents, and year.* [Online]. Available from: https://www.ssb.no/en/statbank/table/11342/tableViewLayout1/ (downloaded 31st May 2024).

[2] Norwegian Institute of Public Health (2024). *Youth profiles for municipalities and districts.* [Online]. Available from: https://www.fhi.no/op/oppvekstprofiler/ (downloaded 31st May 2024)

**Figure S2.** Overview of Trysil municipality and the location of the activity space.

**Target group, location, and aim**

Trysil municipality aims to develop an activity space for youth on the flood embankment in Innbygda, the main center of Trysil, which hosts close to 40% of the population in the municipality. See Figure S2 for a general overview of the municipality.

The activity space aims to connect the main center with the river passing by providing an exciting area focusing on physical activity for the youth staying in and visiting the main center. It will also be established zones to sit down and socialize.

**Concept and design elements**

To achieve the aim a solid steel pipeline along the flood embankment will be constructed, creating an obstacle course. The foundations of the pipeline are dressed in various wooden boxes that can be used both as benches and as parkour elements. Through signs and description, the users will be challenged to finish the obstacle course in one stretch. A new beach and stones in the river will be established, providing an opportunity to swim and wade. The flow of water in the river determines how many steppingstones appear. Other main elements are several semi-open huts with campfires, and other places to socialize and chill like hammocks and nets.

**Stakeholders involved in the project**

Interdisciplinary municipal project group, local youth, representatives for the development project of the main center, external consultants, and the inhabitants in general.

#### Supplementary information S3. The pool park in Gjøvik

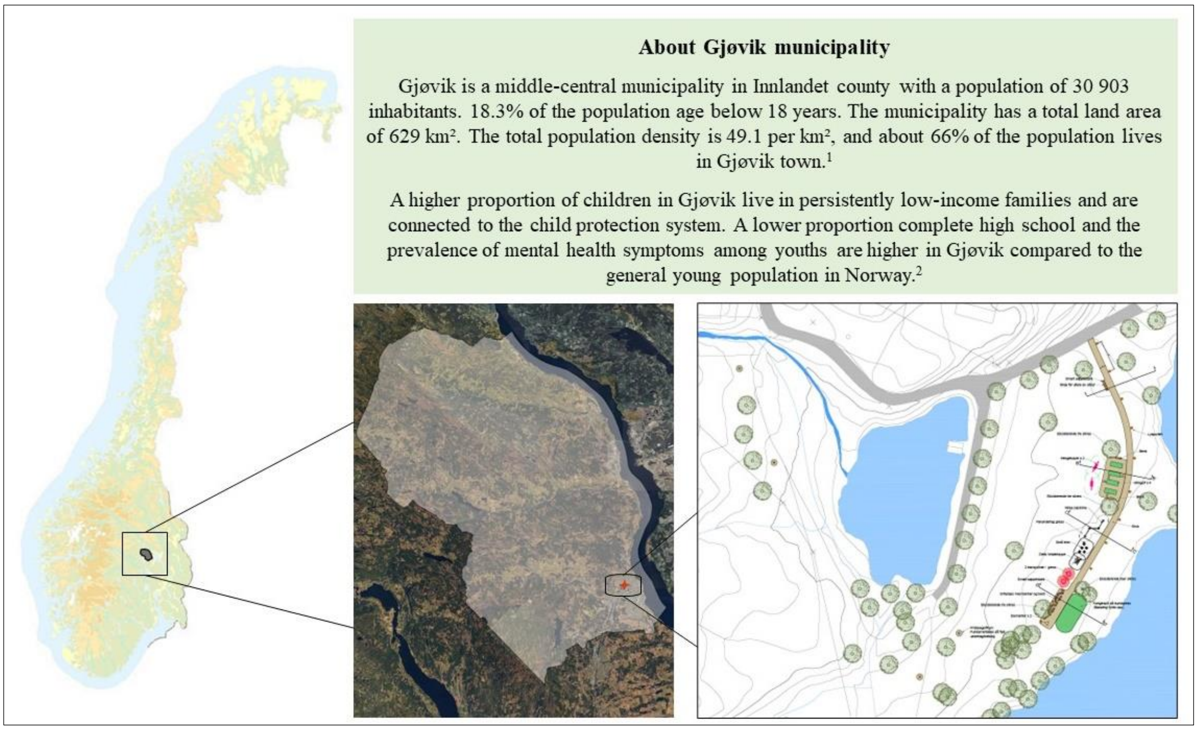


[1] Statistics Norway (2024). *11342: Population and area, by region, contents, and year.* [Online].
Available from: https://www.ssb.no/en/statbank/table/11342/tableViewLayout1/ (downloaded 31st May 2024).
[2] Norwegian Institute of Public Health (2024). *Youth profiles for municipalities and districts.* [Online].
Available from: https://www.fhi.no/op/oppvekstprofiler/ (downloaded 31st May 2024)

**Figure S3.** Overview of Gjøvik municipality and the location of the activity space.

**Target group, location, and aim**
Gjøvik municipality aims to develop an activity space for youth, especially girls. The activity space will be developed just outside Gjøvik town, which hosts two-thirds of the population in the municipality. See Figure S3 for a general overview of the municipality.

The activity space aims to support feelings of relaxation, happiness, and safety. It is supposed to be a space for having fun and for being social. The activity space should provide a warm atmosphere with predominantly low-intensity activities. These activities do not necessitate traditional sports skills and allow users to participate individually or together with others.

**Concept and design elements**
To achieve the aim the activity space will be divided into four zones. Zone 1: Break room/ hanging area. Designed as a place for relaxation and socialization with benches and barbeques. Zone 2: Chill and something challenging. A cool hangout, where you can watch others' activities. Net between the trees to hang and climb in, and other design elements challenging balance and flexibility. Zone 3: Low heart rate. Activities where precision and eye-hand coordination are in focus, like Mini golf, Boccia, and Frisbee golf. Zone 4: Fine motor skills and collaboration. Activities you can do together that require focus and fine motor skills. In addition to the activities and seating, a terminal will be established for self-service of equipment.

**Stakeholders involved in the project**
Interdisciplinary municipal project group, local youths, users of and staff at the child and adolescent psychiatric clinic, local history and theater group, scout association, external consultants/landscape architects.

### Supplementary information S4. The Parcade in Larvik

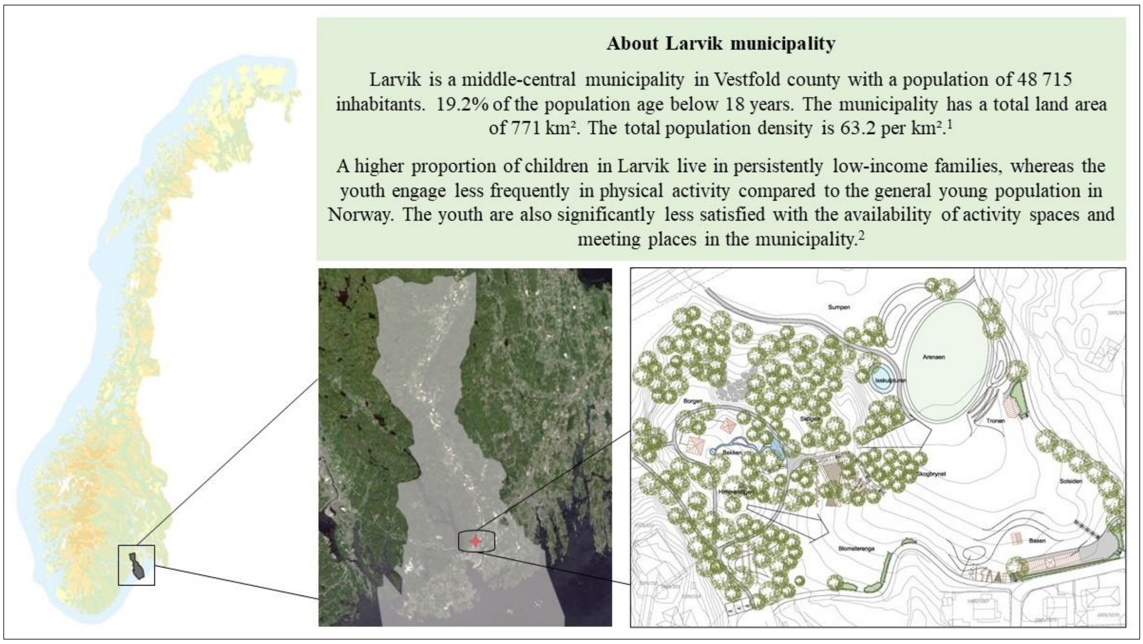


[1] Statistics Norway (2024). *11342: Population and area, by region, contents, and year.* [Online].
Available from: https://www.ssb.no/en/statbank/table/11342/tableViewLayout1/ (downloaded 31st May 2024).
[2] Norwegian Institute of Public Health (2023). *Youth profiles for municipalities and districts.* [Online].
Available from: https://www.fhi.no/op/oppvekstprofiler/ (downloaded 31st May 2024)

**Figure S4.** Overview of Larvik municipality and the location of the activity space.

**Target group, location, and aim**
Larvik municipality aims to develop a year-round activity space for youth. The activity space will be located just outside Larvik city centre, which hosts slightly above 50% of the population in the municipality. See Figure S4 for a general overview of the municipality.

The activity space will provide a vibrant environment where youth can engage in physical, social, and digital activities. By integrating natural qualities and technology, the Parcade will offer a dynamic arena for movement and interaction through game-based activities. The space will be designed for social interaction and physical challenges, mirroring popular video games, and requiring a physical presence in the park to participate in.

**Concept and design elements**
To achieve the aim, several exciting and diverse game and activity zones will be created, including a mobile app where youth can create their own avatars. Over five construction stages, twelve zones will be developed, including “the arena”, featuring various ball fields, workout furniture, and interactive play equipment, as well an amphitheatre stage and a DJ booth. In “the castle” zone, youth can ascend to the top of a viewing tower via ramps and stairs, with a fireman's pole for a speedy decent. “The slide and steps” zone offers a semi-open slide with integrated light and sounds effects, steps on both side, and competitive elements accessible through the app. The design elements will incorporate local materials and shapes reminiscent of popular video games, harmonizing with the landscape. In the park, young gamers can earn “parcoins” by participating in physical challenges across different zones. The physical elements will be designed to function both with and without gaming.

**Stakeholders involved in the project**
Interdisciplinary municipal project group, youth, youth gaming group, local housing and welfare association external consultants/landscape architects, game developers and designers.

### Supplementary information S5. The Bruer park in Vestby

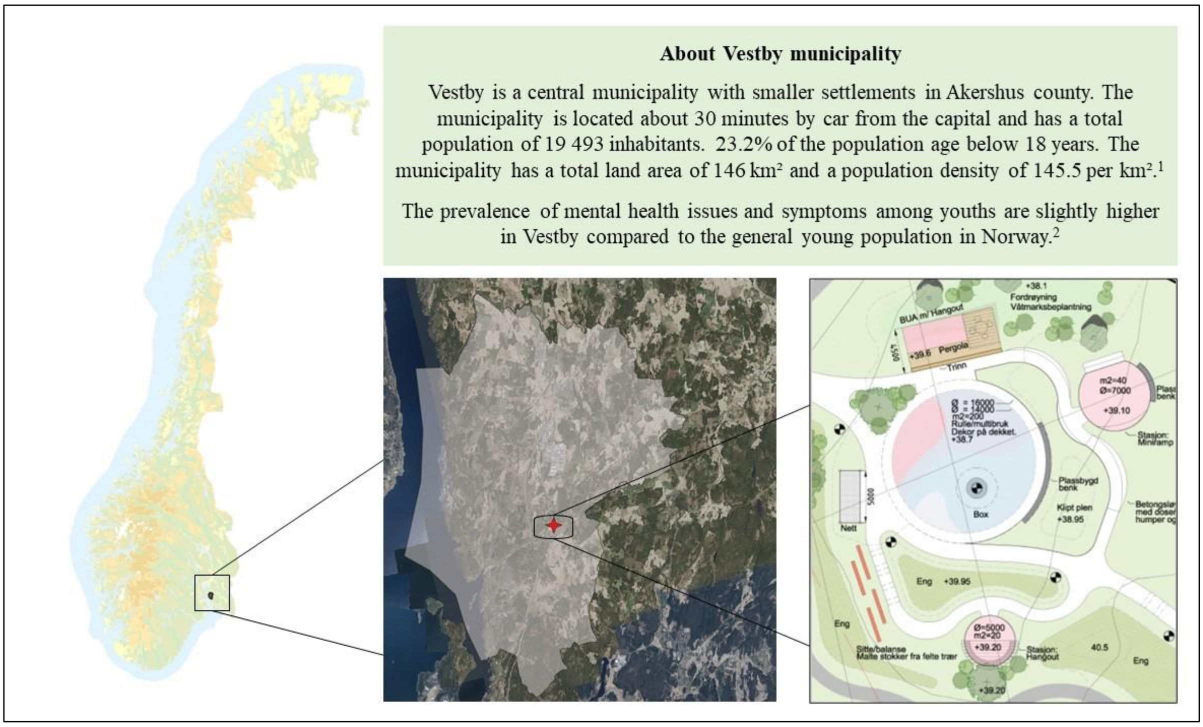


[1] Statistics Norway (2024). *11342: Population and area, by region, contents, and year.* [Online]. Available from: https://www.ssb.no/en/statbank/table/11342/tableViewLayout1/ (downloaded 31st May 2024).
[2] Norwegian Institute of Public Health (2023). *Youth profiles for municipalities and districts.* [Online]. Available from: https://www.fhi.no/op/oppvekstprofiler/ (downloaded 31st May 2024)

**Figure S5.** Overview of Vestby municipality and the location of the activity space.

**Target group, location, and aim**

Vestby municipality aims to develop an activity space for youth, with a special focus on girls. The activity space will be developed in the local center Hølen, which hosts about 6% of the population in the municipality. See Figure S5 for a general overview of the municipality.

The goal is to establish an attractive, social, and inclusive activity space for girls who want to roller skate. The area will also cater to beginners and younger children engaged in various types of wheel-based activities. Other user groups include individuals passing through via the new rolling and walking path along the old Smaalensbanen train track.

**Concept and design elements**

To achieve the aim, a circular roller-skating arena with a 16-meter diameter concrete surface will be established. Surrounding the arena, asphalt paths will be constructed in shapes like rolling waves, including two circle shaped stations with mini ramps and benches. The stations will be specifically designed for beginners to practice in a comfortable environment. The roller-skating arena and its surroundings will be decorated with diverse artistic elements. Adjacent to the roller-skating arena, a container with glass windows will serve as a self-service equipment shed, lending free of charge roller skating equipment. Tables and benches will be places outside the shed. Other hangout areas will be located along the edge of the roller-skating arena, including a large net and tree trunks along the path.

**Stakeholders involved in the project**

Interdisciplinary municipal project group, local girls (age 11-19), Young arena (low threshold offer for youth in Vestby), Girls on Wheels (a roller-skating club), and external consultants/landscape architects.

#### Supplementary information S6. Little Venice of Norway, the Siggerud canal

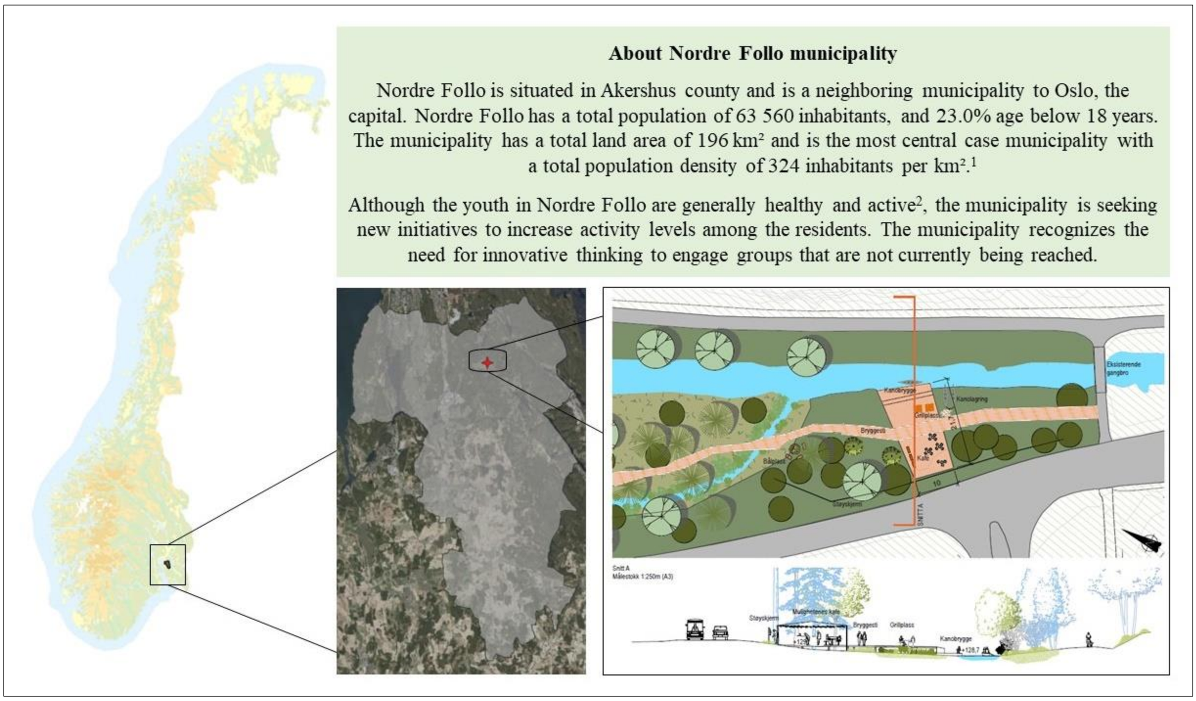


[1] Statistics Norway (2024). *11342: Population and area, by region, contents, and year.* [Online]. Available from: https://www.ssb.no/en/statbank/table/11342/tableViewLayout1/ (downloaded 31st May 2024).
[2] Norwegian Institute of Public Health (2023). *Youth profiles for municipalities and districts.* [Online]. Available from: https://www.fhi.no/op/oppvekstprofiler/ (downloaded 31st May 2024)

**Figure S6.** Overview of Nordre Follo municipality and the location of the activity space.

**Target group, location, and aim**
Nordre Follo will develop a year-round activity space along parts of the Siggerud canal (2-3 km in length) that is intended for everyone, but with a special focus on youth. The activity space will traverse the local center of Siggerud and serve as a local attraction and a regional destination. See Figure S6 for a general overview of the municipality.

The aim is to create an activity space that harmonizes with nature and connects the canal to the nearby lake (Langen), which currently serves as a regional destination with canoeing in the summer and ice-skating in the winter.

**Concept and design elements**
To achieve the aim, two "harbours" will be established at each end of the canal, offering free canoeing equipment, pedal boats, and SUPs. One of the harbours will house the "Café of possibilities" featuring a simple roof, outdoor furniture, and grills. The café will provide space for sustainable and creative activities, such as swapping or selling used clothes or books, as well as temporary shops (popup shops) for fundraising purposes. A canal path will also be constructed, adorned with small information signs about the nature, plant, and animal life in the area. Throughout the year, the water level in the canal will fluctuate, providing varying conditions for those interested in paddling or other water activities. A muscle powered raft will be constructed to cross the canal. When using the raft, local youth can enjoy a socialization zone with a semi-open hut with benches and hammocks.

**Stakeholders involved in the project**
Interdisciplinary municipal project group, local youth, interest groups and volunteers, the public, and external consultants.
